# Plasma Metallomics Reveals Potential Biomarkers and Insights into the Ambivalent Associations of Elements with Acute Myocardial Infarction

**DOI:** 10.1101/2022.07.14.22277628

**Authors:** Si Ying Lim, Hiranya Dayal, Song Jie Seah, Regina Pei Woon Tan, Zhi En Low, Anna Karen Carrasco Laserna, Sock Hwee Tan, Mark Y. Chan, Sam Fong Yau Li

## Abstract

Acute myocardial infarction (AMI) is a leading cause of mortality and morbidity worldwide. Using a validated and efficient ICP-MS/MS-based workflow, a total of 30 metallomic features were profiled in a study comprising 101 AMI patients and 66 age-matched healthy controls. The metallomic features include 12 essential elements (Ca, Co, Cu, Fe, K, Mg, Mn, Na, P, S, Se, Zn), 8 non-essential/toxic elements (Al, As, Ba, Cd, Cr, Ni, Rb, Sr, U, V), and 10 clinically relevant element-pair product/ratios (Ca/Mg, Ca×P, Cu/Se, Cu/Zn, Fe/Cu, P/Mg, Na/K, Zn/Se). Preliminary linear regression with feature selection confirmed smoking status as a predominant determinant for the non-essential/toxic elements, and revealed potential routes of action. Univariate assessments with adjustments for covariates revealed insights into the ambivalent relationships of Cu, Fe, and P with AMI, while also confirming cardioprotective associations of Se. Also, beyond their roles as risk factors, Cu and Se may be involved in the response mechanism in AMI onset/intervention, as demonstrated via longitudinal data analysis with 2 additional time-points (1-/6-month follow-up). Finally, based on both univariate tests and multivariate classification modelling, potentially more sensitive markers measured as element-pair ratios were identified (e.g., Cu/Se, Fe/Cu). Overall, metallomics-based biomarkers may have utility for AMI prediction.

## 1. Introduction

Acute myocardial infarction (AMI), a condition classified under coronary heart disease (CHD), is one of the leading causes of mortality and morbidity worldwide despite remarkable progress in disease diagnosis and treatment.^1 2^ Both clinical and epidemiological studies have demonstrated the cardioprotective and detrimental associations of essential minerals and non-essential/toxic metal exposure respectively, with risks of AMI, CHD, or cardiovascular disease (CVD) in general.^3, 4^ However, given the global-acting and non-independent nature of these various metals in health and disease, their values as biomarkers are limited. Moreover, the benefits of public health efforts and intervention strategies (*e*.*g*., via dietary supplementation or reducing anthropogenic exposure) remain unclarified.^3, 4^ Thus, more accurate and in-depth characterisation of the associations between these different types of elements with AMI aetiology is vital.

Thus, omics strategies such as metallomics have great merit, as the interplay between various non-independent elements and other biomolecules may be revealed. However, as shown in a recent bibliometric analysis on omics applications in CHD research, comprehensive metallomics analysis has seldom been done in this area, and there was only 1 study on blood plasma metallomics study in a subgroup of AMI patients.^5^ This could be because metals have rarely been conjugated with the omics world unlike the other omics frontrunners, and that metallomics only garnered greater attention recently.^5, 6^ Nevertheless, metallomics has already been widely applied in environmental studies and toxicological appraisals, and methods have been rapidly developed to enable quick and efficient metallomic profiling.^7^ This study’s overarching goal is therefore to capitalise on such developments for bridging existing research gaps regarding the AMI metallome.

With that overarching goal, our study aims to assess both essential elements and non-essential/toxic elements with as wide a coverage as possible. To achieve that, the inductively coupled plasma – mass spectrometry (ICP-MS) with MS/MS and collision/reaction cell (CRC) capabilities was used for its remarkable sensitivity, enabling the analysis of more elements at trace and ultra-trace levels.^8^ Also, beyond solely comparing elemental concentrations in the blood plasma, element ratios are also relevant. This is because the dynamic balance or metabolic crossroads of multiple elements have implications on various biological processes in human health and disease, and could also potentially link health/disease with diet and environmental changes.^9^ Thus, we identified a few element-pairs relevant to CVD and in human health in general: Ca/Mg^10, 11^, Ca×P^12-14^, Cu/Se^15^, Cu/Zn^16, 17^, Fe/Cu^18^, Na/K^19, 20^, P/Mg^21^, and Zn/Se^15^. Our work thereby includes these additional variables as part of the metallomics profile, to paint a more comprehensive picture about these elements in AMI.

In summary, because few studies have examined the association between circulating element (both essential and non-essential/toxic) levels and AMI, and none so far has also comprehensively assessed several element product/ratios in Asian populations across 3 time-points, our study presents useful insights into the pertinent disturbances in AMI metallome.

## 2. Materials and methods

### 2.1. Reagents and chemicals

All chemicals used were of trace analysis grade (99.999%) unless otherwise stated, and ultrapure water was also used for all chemical and sample preparation. The multi-element standard (IV-ICPMS-71A) was purchased from Inorganic Ventures (Christiansburg, VA, USA), and it contains the following elements in 3% nitric acid: Ag, Al, As, B, Ba, Be, Ca, Cd, Ce, Co, Cr, Cs, Cu, Dy, Er, Eu, Fe, Ga, Gd, Ho, K, La, Lu, Mg, Mn, Na, Nd, Ni, P, Pb, Pr, Rb, S, Se, Sm, Sr, Th, Ti, Tm, U, V, Yb, Zn. The ICPMS internal standard mix was purchased from Agilent Technologies (Santa Clara, CA, USA, and it contains Bi, Ge, In, Li-6, Rh, Sc and Tb. The 60% nitric acid (ultrapure; any metals at sub ppt levels) used for all experiments was purchased from Kanto Chemical Co. (Tokyo, Japan).

### 2.2. Patients and controls

This study was approved by the National Healthcare Group Doman Specific Review Board (NHG DSRB; REF NO. 2013-00248 and 2016-00210), and all subjects gave their informed consent to participate prior to the inclusion. All experiments were performed in compliance with the relevant laws and institutional guidelines. Blood plasma samples from 101 AMI patients were collected from the National University Heart Centre, Tan Tock Seng Hospital, Changi General Hospital, Sarawak General Hospital Heart Centre, and Christchurch District Hospital. AMI plasma samples were collected 24-48 hours post-percutaneous coronary intervention (PCI). Samples from two additional follow-up time-points were also collected from the AMI patients (T1: 24 – 48 hours post-PCI; T2: 1 month after T1; T3: 6 months after T1). Blood plasma samples from 66 matched healthy community-dwelling controls were collected from Singapore. All blood samples were immediately centrifuged at – 4 °C and were stored at – 80 °C prior to analysis.

### 2.3. Metallomics sample preparation

To obtain elemental samples from human blood plasma, acid digestion aided with heating via a hotplate was done. Specifically, 80 μL blood plasma was added into a 15 mL polypropylene centrifuge tube, followed by the addition of 80 μL ultrapure water and 80 μL ultrapure nitric acid. The centrifuge tubes were gently swirled to allow mixing. Immediately after, the samples were heated using a hotplate at 98 °C for 2 hours. To further process the samples for ICP-MS analysis, the digested samples were diluted to 8 mL with ultrapure water. The diluted samples were then filtered using 2.2 μm PTFE syringe filters into clean centrifuge tubes. Blank samples were prepared in the same way, but with the blood plasma component replaced with ultrapure water. The samples were then stored at 4 °C until ICP-MS analysis as recommended by a recent report.^22^

### 2.4. ICP-MS analysis

ICP-MS analysis was done using the 8900 ICP-MS/MS system (equipped with triple quadruple mass spectrometer) from Agilent Technologies. To optimise sensitivity and accuracy, various CRC modes using the tandem mass spectrometer were used to quantitate different elements. Supplementary Table S1 summarises the instrument and CRC parameters. To correct for matrix effects and non-spectral interferences, multiple elements from the internal standard mix were spiked online and elemental concentrations were normalised to the internal standards. To obtain optimal precision and accuracy, the internal standard analyte for each element was selected as close in mass number as possible to that of the analyte element, with ionisation energies considered as well. The masses and mass shifts used for integration for each element and the corresponding internal standard analytes are included in Supplementary Table S2. To obtain elemental concentrations of the samples, external standard calibration was done using the multi-element standard prepared in various concentrations. It was noted that a few elements (Al, Ca, K, Mg, Na, P, S) were present in higher abundance where their concentrations in some or all of the plasma samples exceeds the external standard calibration range. To determine their concentrations, the samples were diluted (dilution factor = 50) and re-analysed by the ICP-MS.

### 2.5. Data processing and statistical analysis

All statistical analyses were conducted in *R* (R Core Team).^23^ Normality of all metabolite and glycan features was first assessed via the Kolmorov-Smirnov test. Since non-normal distributions (p < 0.05 based on Kolmorov-Smirnov test) were found with some of the features, non-parametric tests were done subsequently. Baseline characteristics among patients and controls were compared using Mann Whitney test for continuous variables, and Fisher’s Exact test for categorical variables. Differences in plasma levels of the metallomic features between patient groups were also assessed using the Mann Whitney test. Linear regression modelling for identifying independent determinants of the metallomic features amongst covariables was done with backwards elimination to remove non-significant variables until all variables remaining in the model had *p-*value < 0.05. Analysis of covariance (ANCOVA) with the inclusion of significant determinants as covariates was done by implementing a generalised linear model (GLM) with Bonferroni correction for multiple comparisons. Analysis of the longitudinal data with 3 time-points was achieved via Friedman test, and Kendall’s W coefficient of concordance was evaluated as the effect size for Friedman test.

Classification modelling was done on MetaboAnalyst (http://www.metaboanalyst.ca). To prepare the dataset with both clinical and metallomics variables for multivariate analysis, log-transformation and scaling via mean-centring was done. The receiving operating characteristic (ROC) curve for biomarker identification and performance evaluation was generated based on random forest classification modelling with repeated random sub-sampling cross-validation (30 iterations; each iteration uses 2/3 samples for feature selection and model training, and remaining 1/3 for testing). Important features were identified and ranked based on decreases in accuracy.

## 3. Results and discussion

### 3.1. Generation of plasma metallomic profiles

Out of 43 elements from the multi-element standard, 22 were successfully quantified from the human blood plasma samples via ICP-MS analysis as described. To validate that the sample preparation workflow involving nitric acid digestion with hotplate-assisted heating was sufficient to digest and stabilise the 22 detected elements, and that the CRC modes and internal standard elements were appropriate, method repeatability and recovery were assessed.

For assessing intra-day repeatability/precision, three independent digestions of the same pooled plasma sample spiked with multi-element standard (final concentration of each element = 500 ppb) were done and analysed. Additional tests with dilution (dilution factor = 50) were also done for evaluating some of the elements present at lower amounts in blood plasma. For assessing inter-day repeatability or intermediate precision, nine independent extractions from the same pooled plasma sample were prepared over three days. All replicates were analysed using the ICP-MS using the same operating conditions. RSD values (both intra- and inter-day) for all 22 elements were low (range: 1.3 – 7.9 %), demonstrating satisfactory method repeatability/precision. The RSD values are deemed satisfactory by cross-checking with recent publications on ICP-QQQ-based metallomics methods.^24, 25^ Next, to assess recovery, element concentrations obtained from samples spiked with multi-element standard before and after digestion were compared. All 22 elements have recoveries roughly within +/-10 % threshold from 100 % (range: 89.9 – 105.2 %). This indicates satisfactory recovery of the entire analytical workflow. Additionally, it was also confirmed in this study that all elements analysed have baseline equivalent concentrations lower than their quantified concentrations in the pooled plasma sample, and that the calibration curves have R^2^ values > 0.9995 which demonstrates good linearity.

However, we note that there were also some biologically relevant elements not quantifiable using our analytical workflow. Prominent examples include mercury as well as halogens like iodine, where additional steps and reagents may be required to counter issues with stability, memory effects, etc.^26, 27^ Nonetheless, together with the satisfactory precision, recovery, and good analyte coverage of 22 elements, this metallomics workflow without the need for microwave digestion was simple and efficient. Thus, we applied it on the plasma samples of AMI patients and healthy controls to generate their metallomic profiles first consisting of elemental concentration data. Relevant element-pair product/ratios were also calculated. The 12 essential elements studied here are Ca, Co, Cu, Fe, K, Mg, Mn, Na, P, S, Se, and Zn. The 10 non-essential or toxic elements are Al, As, Ba, Cd, Cr, Ni, Rb, Sr, U, V. As a whole, metallomic profiles comprising both elemental concentrations and element-pair product or ratios (30 metallomic features in total) were generated for each sample.

### 3.2. Baseline characteristics and their associations with the metallome

The baseline characteristics (including demographic characteristics, medical history related to the disease, as well as traditional cardiovascular risk factors and predictors of AMI and adverse events) of the study populations are summarised in Table 1. As the healthy controls were age-matched to the AMI cases, there are no statistically significant differences (*p* > 0.05) in age distributions between the AMI and healthy groups. However, the distributions of gender, smoking status, and race were found to be significantly different between the two groups (*p* < 0.0001). Next, for the medical history (Table 1), they are closely matched between AMI and control groups, with the only exception being dyslipidemia (*p* < 0.001). For variables such as troponin T as measured via the high-sensitivity test (hsTnT) and N-terminal-pro hormone brain natriuretic peptides (NTproBNP), the highly significant differences are expected as they are biomarkers of AMI, and positive troponin was also an inclusion criterion for the AMI cases. As for other traditional cardiovascular risk factors, several of them (diastolic blood pressure (DBP), systolic blood pressure (SBP), high-density lipoprotein-cholesterol (HDL-C), white blood cell count (WBC), and creatinine levels are also statistically significant variables (*p* < 0.0001).

**Table 1.** Baseline characteristics of the study populations.

| Characteristic | AMI (n=101) | Control (n=66) | p-value<sup>a</sup> |
| --- | --- | --- | --- |
| ***Demographic variables*** |  |  |  |
| Age (Median [Min-Max]) | 57<br>(33 - 81) | 54.5<br>(40 - 71) | 2.66E-01 |
| Gender (Female) (%) | 12.9 | 40.9 | <u>6.30E-04</u> |
| Smoker (%) |  |  |  |
| Non | 37.6 | 90.9 |  |
| Current | 35.6 | 3.0 |  |
| Ex | 26.7 | 6.1 |  |
| Race (%) |  |  | <u>1.00E-14</u> |
| Chinese | 27.7 | 60.6 |  |
| Malay | 21.8 | 9.1 |  |
| Indian | 6.9 | 30.3 |  |
| Others | 43.6 | 0.0 |  |
| ***Cardiovascular risk factors based on medical history*** |  |  |  |
| Prior MI (%) | 4.0 | 0.0 | 1.54E-01 |
| FH of CHD (%) | 20.8 | 10.6 | 9.40E-02 |
| Diabetes (%) | 20.8 | 12.1 | 2.10E-01 |
| Hypertension (%) | 43.6 | 28.8 | 7.20E-02 |
| Dyslipidemia (%) | 42.6 | 77.3 | <u>3.30E-05</u> |
| ***Other cardiovascular risk factors, predictors of AMI and adverse events (Median [Min-Max])*** |  |  |  |
| DBP, mm Hg | 83.50<br>(39 - 138) | 74<br>(53 - 107) | <u>1.65E-04</u> |
| SBP, mm Hg | 140<br>(74 - 241) | 127<br>(97 - 175) | <u>8.22E-06</u> |
| Triglyceride, mmol/L | 1.50<br>(0.35 - 5.58) | 1.38<br>(0.37 - 4.37) | 5.12E-01 |
| Cholesterol, mmol/L | 5.01<br>(3.20 - 7.40) | 5.32<br>(4.05 - 7.37) | <u>1.51E-03</u> |
| HDL-C, mmol/L | 1.03<br>(0.67 - 9.40) | 1.34<br>(0.80 - 2.31) | <u>6.64E-10</u> |
| LDL-C, mmol/L | 3.20<br>(1.60 - 5.51) | 3.27<br>(2.19 - 5.25) | 1.07E-01 |
| WBC, × 10<sup>9</sup>/L | 10.85<br>(4.40 - 25.90) | 5.50<br>(2.60 - 9.0) | <u>1.46E-20</u> |
| Platelet, × 10<sup>9</sup>/L | 250.5<br>(134 - 649) | 258<br>(165 - 599) | 3.79E-01 |
| Creatinine, mmol/L | 88.5<br>(51 - 142) | 69<br>(42 - 97) | <u>3.90E-13</u> |
| hsTnT, pg/mL | 1726<br>(62 - 9819) | 6<br>(3 - 18) | <u>8.83E-28</u> |
| NTproBNP, pg/mL | 718<br>(68 - 6819) | 36<br>(7 - 150) | <u>3.01E-27</u> |

<sup>a</sup> Significant *p*-values are underlined.
Abbreviations used in the table: FH, family history; DBP, diastolic blood pressure; SBP, systolic blood pressure; HDL-C, high-density lipoprotein cholesterol; LDL-C, low-density lipoprotein cholesterol; WBC, white blood cells; hsTnT, high-sensitivity troponin T; NTproBNP, N-terminal-pro hormone brain natriuretic peptides.

We note that the above variables could be possible confounders in this metallomics study. As such, before comparing the metallomic profiles between the AMI and healthy groups, we determined whether the aforementioned demographic characteristics and traditional CVD risk factors were independent determinants of plasma levels of the metallomic features. For that purpose, we fitted a linear model with each metallomics feature as the dependent variable and the demographic characteristics and traditional CVD risk biomarkers as independent variables.

Backwards elimination was performed to remove non-significant independent variables until all variables remaining in the model had *p-*values < 0.05. The β coefficients and significance levels of the determinants are summarised in Table 2. The determinants identified for each metallomic feature would then be included as covariates for ANCOVA when assessing statistical differences between AMI versus healthy in the next section.

**Table 2.** Independent determinants of each metallomic feature among study participants based on backwards elimination linear regression.

| Metallomic feature | Independent determinant(s)<sup>a</sup> | $\beta^b$ | Standard error | Level of significance<sup>c</sup> |
| --- | --- | --- | --- | --- |
| *Essential elements* |  |  |  |  |
| Ca, µg/mL | nil |  |  |  |
| Co, ng/mL | nil |  |  |  |
| Cu, µg/mL | Gender (Female) | 0.15 | 0.04 | ** |
|  | Cholesterol | -0.12 | 0.04 | ** |
|  | LDL-C | 0.14 | 0.05 | ** |
| Fe, µg/mL | nil |  |  |  |
| K, µg/mL | nil |  |  |  |
| Mg, µg/mL | Race (Indian) | -4.92 | 2.42 | * |
|  | Smoker (Current) | 11.85 | 2.16 | **** |
| Mn, ng/mL | Race (Chinese) | -6.29 | 1.99 | ** |
| Na, mg/mL | nil |  |  |  |
| P, µg/mL | nil |  |  |  |
| S, µg/mL | nil |  |  |  |
| Se, ng/mL | Race (Chinese) | 23.84 | 3.91 | **** |
|  | Race (Malay) | 17.12 | 4.92 | *** |
| Zn, µg/mL | Race (Chinese) | -10.82 | 3.19 | *** |
|  | Race (Indian) | -12.32 | 3.93 | ** |
|  | Smoker (Current) | 13.98 | 3.00 | **** |
| *Non-essential or toxic elements* |  |  |  |  |
| Al, ng/mL | Smoker (Current) | 685.80 | 214.09 | ** |
| As, ng/mL | White blood cell | -0.14 | 0.048 | ** |
| Ba, ng/mL | Race (Indian) | 120.94 | 41.96 | ** |
|  | White blood cell | 11.50 | 3.89 | ** |
| Cd, ng/mL | Smoker (Current) | 0.25 | 0.06 | **** |
| Cr, ng/mL | Race (Indian) | 91.35 | 30.08 | ** |
| Ni, ng/mL | nil |  |  |  |
| Rb, ng/mL | Smoker (Current) | 33.59 | 11.40 | ** |
| Sr, ng/mL | Smoker (Current) | 29.27 | 5.24 | **** |
| U, ng/mL | Smoker (Current) | 0.51 | 0.11 | **** |
| V, ng/mL | Smoker (Current) | 2.10 | 0.50 | **** |
| *Element product or ratios* |  |  |  |  |
| Ca/Mg | Smoker (Current) | -0.035 | 0.011 | ** |
|  | Cholesterol | 0.038 | 0.012 | ** |
|  | LDL-C | -0.032 | 0.013 | * |
| Ca×P, × 10<sup>-2</sup> | Smoker (Current) | 0.977 | 0.361 | ** |
|  | Cholesterol | -0.446 | 0.163 | ** |
|  | Triglyceride | -0.339 | 0.171 | * |
| Cu/Se | Race (Chinese) | -4.100 | 0.551 | **** |
|  | Race (Malay) | -2.808 | 0.689 | **** |
|  | Gender (Female) | 1.604 | 0.534 | ** |
|  | Cholesterol | -1.298 | 0.566 | * |
|  | LDL-C | 1.553 | 0.613 | * |
| Cu/Zn | Creatinine | 0.003 | 0.001 | * |
| Fe/Cu | nil |  |  |  |
| P/Mg | nil |  |  |  |
| Na/K | nil |  |  |  |
| Zn/Se | Smoker (Current) | 127.49 | 26.98 | **** |

<sup>a</sup> Variables presented here are the significant variables ( $\alpha = 0.01$ ) after backwards elimination of model fitted with age, gender, smoking status, race, dyslipidemia, triglyceride, cholesterol, HDL-C, LDL-C, white blood cell, platelet, and creatinine as independent variables.
<sup>b</sup> Difference in mean levels of the respective element or element ratio/product associated with a 1-unit increase in the independent variable. Units for each variable can be found in Table 1.
<sup>c</sup> Level of significance: \*, $p \leq 0.05$ ; \*\*, $p \leq 0.01$ ; \*\*\*, $p \leq 0.001$ ; \*\*\*\*, $p \leq 0.0001$ .

Regardless, with reference to Table 2, various noteworthy associations of metallomic features with demographic characteristics and cardiovascular risk factors were identified. Firstly, smoking status is associated with the greatest number of metallomic features, and predominantly in the non-essential or toxic elements (*i.e*., Al, Cd, Rb, Sr, U, V; all *p* < 0.01). This could be expected as several of these elements are found in tobacco, cigarette paper, filters, and cigarette smoke (except U).^28, 29^ It has also been revealed that the levels of these elements in circulation are affected by smoking, and they are linked to the disturbance of metal homeostasis and the pathogenesis of diseases.^28, 29^ Moreover, out of the essential elements, Zn was also negatively associated with current smokers. This decrease could either be attributed to the link between Zn and nicotinic receptors stimulation (lower Zn levels = cause/predisposition of smoking),^30^ or attributed to increased blood Cd concentration which induces expression of metallothionein binding of both Cd and Zn (lower Zn levels = effect of smoking).^28, 31^ This is particularly relevant to our study on AMI as smoking-induced lowered Zn levels may lead to cellular ageing (oxidative stress) and increases the risk of hypertension.^32^

Secondly, three other pertinent metal-cardiovascular risk factor associations are (1) increasing Cu with lowering of cholesterol and increase in LDL-C, (2) increasing As with lowering of white blood cell levels, and (3) increasing Cu/Zn ratio with increasing creatinine (all *p* < 0.01). For Cu, it has been reported that Cu deficiency is associated with increased total cholesterol.^33^ While the direction of association in our study matches for total cholesterol levels, it was the opposite for LDL-C. Nevertheless, this could imply that an increased LDL-C may not be a true artefact of Cu deficiency, but it could be oxidised LDL instead. This observation could be specific to our study on AMI patients, and the postulation could possibly be explained by Cu’s promotion of LDL-C oxidation.^34, 35^ Next, for As, its negative association with white blood cell levels could be expected as various reports have revealed that arsenicosis or As exposure is associated with the prevalence of leukopenia^36^ since As drives apoptosis in human lymphocytes via mechanisms related to oxidative stress, cytokine production, etc.^37^ This is of relevance as an elevated WBC count has been directly associated with CHD and CVD adverse events in the African-American and White populations.^38^ Thus, while As’s toxicity makes high As levels undesirable, it could also be possible to observe higher levels of As in healthy people than AMI patients due to As’s control of WBC counts. Finally, for the positive association of Cu/Zn ratio with creatinine, it serves to highlight the possible complications with renal issues since creatinine is an indicator of kidney malfunctions. Lower Zn has also previously been found in patients with end-stage renal disease while creatinine was shown to be a determinant of plasma Cu levels, augmenting our hypothesis. ^39^

### 3.3. Statistical comparisons of metallomic profiles between AMI and healthy, and across 3 time-points

Mann Whitney test was first employed to identify metallomic features which are significantly different in AMI patients. Table 3 summarises their distributions, the *p*-values, as well as the direction of change in terms of the composition in AMI patients (measured at T1: 24 – 48 hours post-PCI) compared to the healthy. To reduce bias and assess for the possible attenuating influence of the determinants on each metallomic feature, adjusted *p-*values from ANCOVA were also obtained and are provided in Table 3. To augment conjectures made regarding co-acting elements, Spearman correlation analysis between element levels was also done and results are summarised in a correlation heatmap (Figure 1). For the significantly altered metallomic features, further assessment on their changes across 3 time-points was also done to determine if their dysregulation persists after 1 and 6 months (T2, and T3 respectively). For that purpose, Friedman test (non-parametric version of repeated-measures ANOVA) was applied to the longitudinal data to test for any significant changes (Table 4). Violin plots were also generated to visualise their trajectories (Figure 2). It is noted here that the simpler repeated measures test was used instead of more popular and robust options like the generalised linear mixed-effects model or the growth curve model based on characteristics of our data: (1) small number of time-points which we will assess as discrete time-points, (2) complete and no missing data, (3) simple design, comparison across time-points without assessing additional between/within-subjects factors.

**Table 3.**
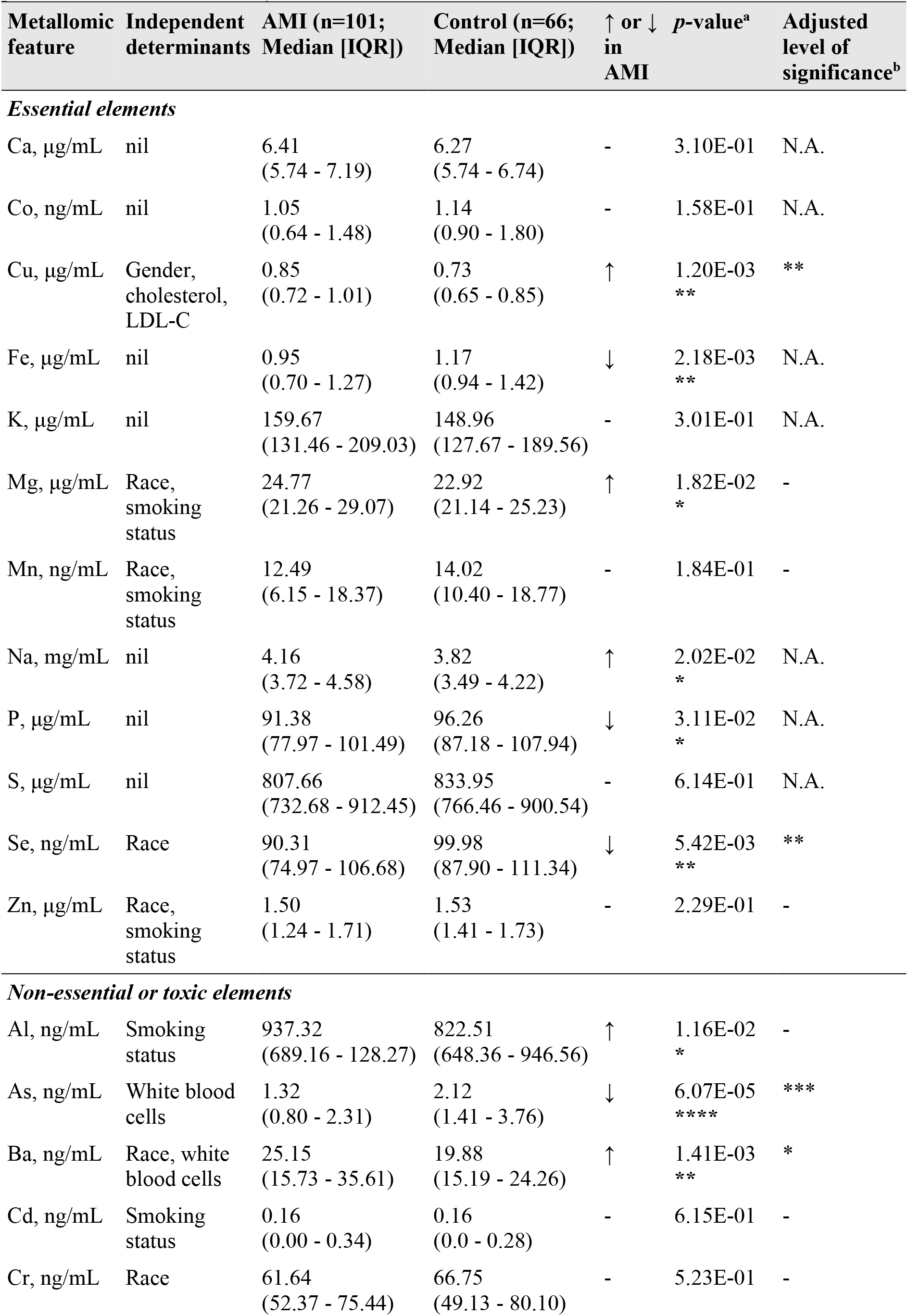
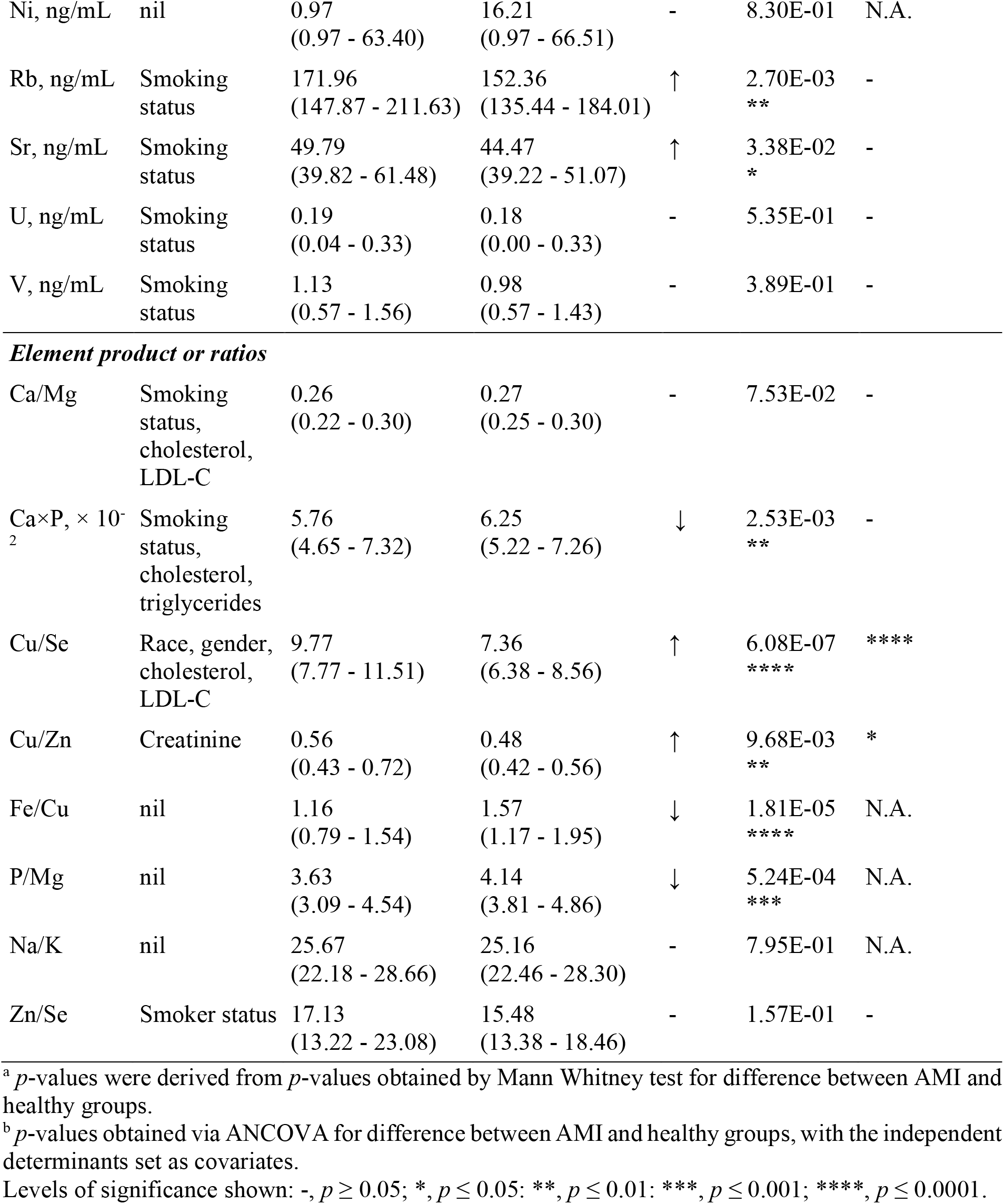
Descriptive statistics and statistical comparisons between metallomic profiles of AMI patients at T1 versus healthy controls.

| Metallomic feature | Independent determinants | AMI (n=101; Median [IQR]) | Control (n=66; Median [IQR]) | ↑ or ↓ in AMI | p-value<sup>a</sup> | Adjusted level of significance<sup>b</sup> |
| --- | --- | --- | --- | --- | --- | --- |
| *Essential elements* |  |  |  |  |  |  |
| Ca, µg/mL | nil | 6.41<br>(5.74 - 7.19) | 6.27<br>(5.74 - 6.74) | - | 3.10E-01 | N.A. |
| Co, ng/mL | nil | 1.05<br>(0.64 - 1.48) | 1.14<br>(0.90 - 1.80) | - | 1.58E-01 | N.A. |
| Cu, µg/mL | Gender, cholesterol, LDL-C | 0.85<br>(0.72 - 1.01) | 0.73<br>(0.65 - 0.85) | ↑ | 1.20E-03<br>** | ** |
| Fe, µg/mL | nil | 0.95<br>(0.70 - 1.27) | 1.17<br>(0.94 - 1.42) | ↓ | 2.18E-03<br>** | N.A. |
| K, µg/mL | nil | 159.67<br>(131.46 - 209.03) | 148.96<br>(127.67 - 189.56) | - | 3.01E-01 | N.A. |
| Mg, µg/mL | Race, smoking status | 24.77<br>(21.26 - 29.07) | 22.92<br>(21.14 - 25.23) | ↑ | 1.82E-02<br>* | - |
| Mn, ng/mL | Race, smoking status | 12.49<br>(6.15 - 18.37) | 14.02<br>(10.40 - 18.77) | - | 1.84E-01 | - |
| Na, mg/mL | nil | 4.16<br>(3.72 - 4.58) | 3.82<br>(3.49 - 4.22) | ↑ | 2.02E-02<br>* | N.A. |
| P, µg/mL | nil | 91.38<br>(77.97 - 101.49) | 96.26<br>(87.18 - 107.94) | ↓ | 3.11E-02<br>* | N.A. |
| S, µg/mL | nil | 807.66<br>(732.68 - 912.45) | 833.95<br>(766.46 - 900.54) | - | 6.14E-01 | N.A. |
| Se, ng/mL | Race | 90.31<br>(74.97 - 106.68) | 99.98<br>(87.90 - 111.34) | ↓ | 5.42E-03<br>** | ** |
| Zn, µg/mL | Race, smoking status | 1.50<br>(1.24 - 1.71) | 1.53<br>(1.41 - 1.73) | - | 2.29E-01 | - |
| *Non-essential or toxic elements* |  |  |  |  |  |  |
| Al, ng/mL | Smoking status | 937.32<br>(689.16 - 128.27) | 822.51<br>(648.36 - 946.56) | ↑ | 1.16E-02<br>* | - |
| As, ng/mL | White blood cells | 1.32<br>(0.80 - 2.31) | 2.12<br>(1.41 - 3.76) | ↓ | 6.07E-05<br>**** | *** |
| Ba, ng/mL | Race, white blood cells | 25.15<br>(15.73 - 35.61) | 19.88<br>(15.19 - 24.26) | ↑ | 1.41E-03<br>** | * |
| Cd, ng/mL | Smoking status | 0.16<br>(0.00 - 0.34) | 0.16<br>(0.0 - 0.28) | - | 6.15E-01 | - |
| Cr, ng/mL | Race | 61.64<br>(52.37 - 75.44) | 66.75<br>(49.13 - 80.10) | - | 5.23E-01 | - |
| Ni, ng/mL | nil | 0.97<br>(0.97 - 63.40) | 16.21<br>(0.97 - 66.51) | - | 8.30E-01 | N.A. |
| Rb, ng/mL | Smoking<br>status | 171.96<br>(147.87 - 211.63) | 152.36<br>(135.44 - 184.01) | ↑ | 2.70E-03<br>** | - |
| Sr, ng/mL | Smoking<br>status | 49.79<br>(39.82 - 61.48) | 44.47<br>(39.22 - 51.07) | ↑ | 3.38E-02<br>* | - |
| U, ng/mL | Smoking<br>status | 0.19<br>(0.04 - 0.33) | 0.18<br>(0.00 - 0.33) | - | 5.35E-01 | - |
| V, ng/mL | Smoking<br>status | 1.13<br>(0.57 - 1.56) | 0.98<br>(0.57 - 1.43) | - | 3.89E-01 | - |
| ***Element product or ratios*** |  |  |  |  |  |  |
| Ca/Mg | Smoking<br>status,<br>cholesterol,<br>LDL-C | 0.26<br>(0.22 - 0.30) | 0.27<br>(0.25 - 0.30) | - | 7.53E-02 | - |
| Ca×P, × 10<sup>-2</sup> | Smoking<br>status,<br>cholesterol,<br>triglycerides | 5.76<br>(4.65 - 7.32) | 6.25<br>(5.22 - 7.26) | ↓ | 2.53E-03<br>** | - |
| Cu/Se | Race, gender,<br>cholesterol,<br>LDL-C | 9.77<br>(7.77 - 11.51) | 7.36<br>(6.38 - 8.56) | ↑ | 6.08E-07<br>**** | **** |
| Cu/Zn | Creatinine | 0.56<br>(0.43 - 0.72) | 0.48<br>(0.42 - 0.56) | ↑ | 9.68E-03<br>** | * |
| Fe/Cu | nil | 1.16<br>(0.79 - 1.54) | 1.57<br>(1.17 - 1.95) | ↓ | 1.81E-05<br>**** | N.A. |
| P/Mg | nil | 3.63<br>(3.09 - 4.54) | 4.14<br>(3.81 - 4.86) | ↓ | 5.24E-04<br>*** | N.A. |
| Na/K | nil | 25.67<br>(22.18 - 28.66) | 25.16<br>(22.46 - 28.30) | - | 7.95E-01 | N.A. |
| Zn/Se | Smoker status | 17.13<br>(13.22 - 23.08) | 15.48<br>(13.38 - 18.46) | - | 1.57E-01 | - |

<sup>a</sup> *p*-values were derived from *p*-values obtained by Mann Whitney test for difference between AMI and healthy groups.
<sup>b</sup> *p*-values obtained via ANCOVA for difference between AMI and healthy groups, with the independent determinants set as covariates.
Levels of significance shown: -, *p* ≥ 0.05; \*, *p* ≤ 0.05; \*\*, *p* ≤ 0.01; \*\*\*, *p* ≤ 0.001; \*\*\*\*, *p* ≤ 0.0001.

**Table 4.**
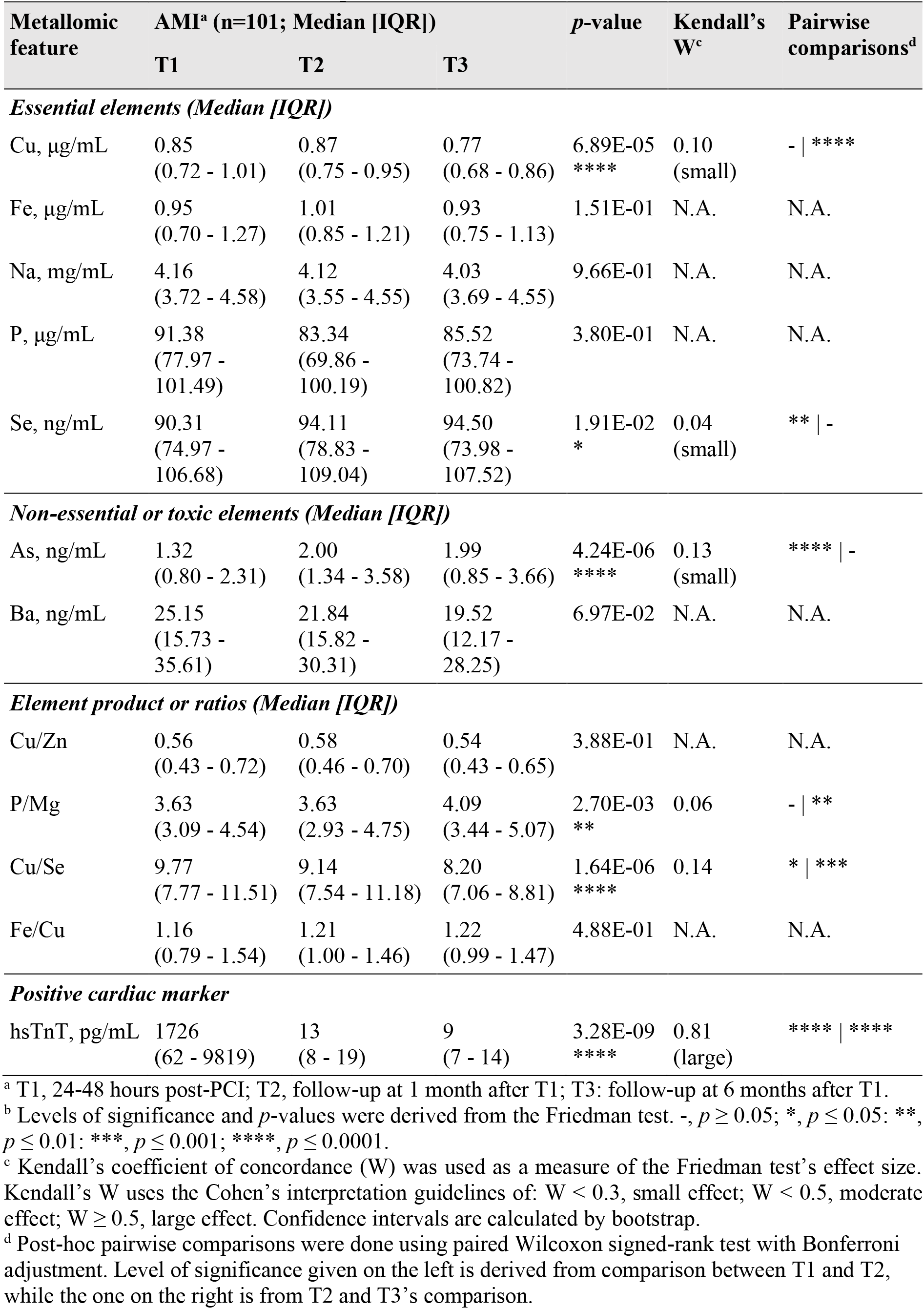
Descriptive statistics and repeated measures ANOVA assessing changes in significant metallomic features across 3 time-points.

**Figure 1.**
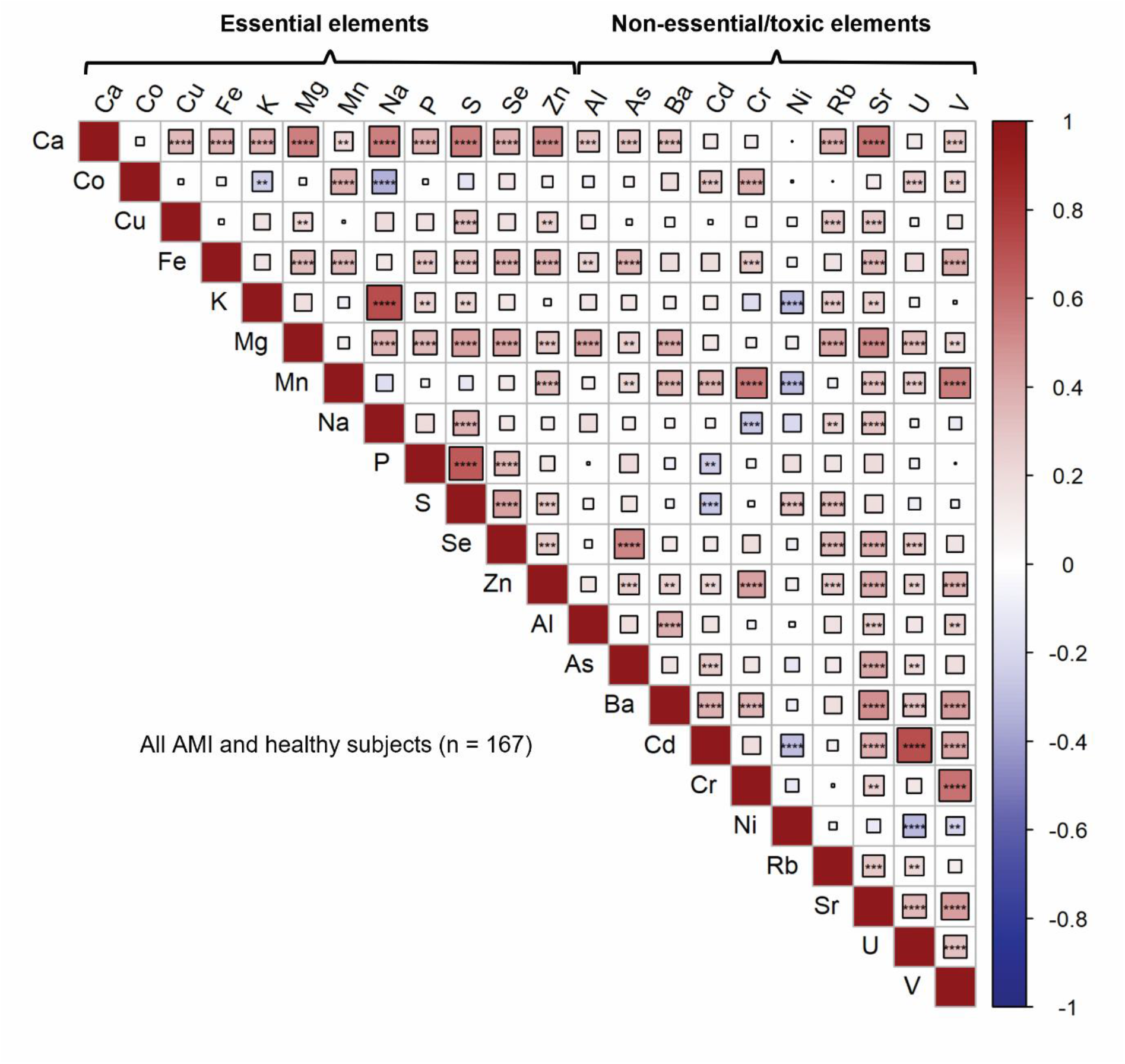
Heatmap showing the correlation between metallomic features (*, *p* ≤ 0.05; **, *p* ≤ 0.01, ***, *p* ≤ 0.001; ****, *p* ≤ 0.0001).

**Figure 2.**
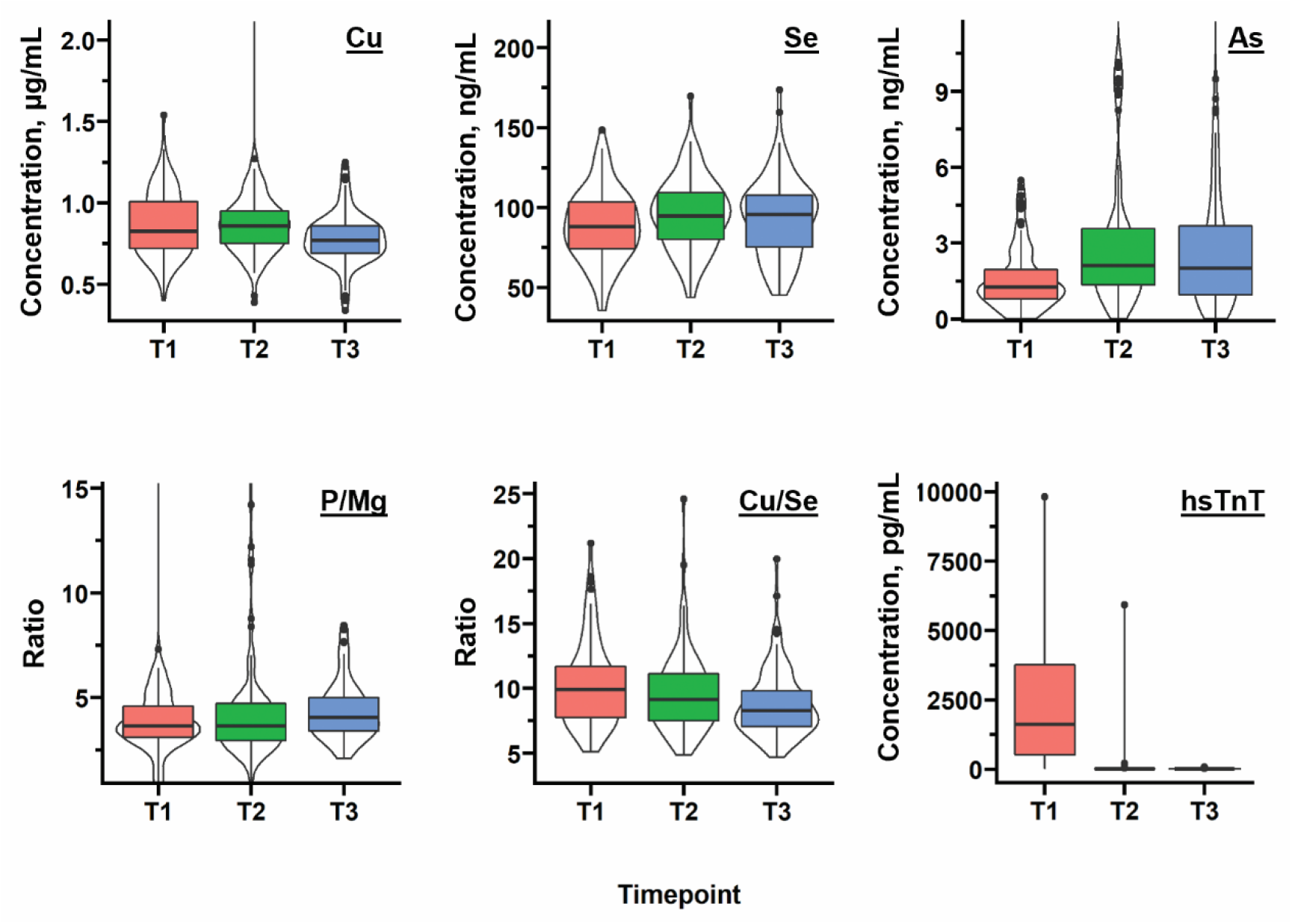
Violin plots showing trajectories of metallomic features with significance changes and hsTnT across 3 time-points (T1, 24-48 hours after PCI; T2, 1 month after T1; T3, 6 months after T1).

#### 3.3.1. Ambivalent relationships of essential element levels with AMI

As a whole, out of the 12 essential elements assessed using the Mann Whitney test, the AMI patients had significantly elevated Cu, Mg, and Na levels in their blood plasma as compared to the healthy controls, while Fe, P, and Se were decreased in levels (all *p* < 0.05; Table 3). However, the *p*-value for Mg was attenuated to a non-significant level when ANCOVA was done. We therefore also exclude Mg as a significant variable for AMI risk. For the rest of the essential elements, significant differences in distributions are expected. Also, out of these essential elements, Cu and Se levels were significantly different at the different time-points (*p* < 0.0001 and 0.05 respectively; Table 4), although detected effect sizes were considered small (0.10 and 0.04 respectively). The possible trajectory of hsTnT, a more specific and choicer biomarker of AMI, is included in Figure 4 for comparison purposes. Since troponin T are released because of cardiac injury and myocardial necrosis (*i.e*., consequence of AMI instead of cause), their serum levels peak 24-48 hours and return to baseline over 10-14 days.^40^ Their trajectory of drastic decrease at T2 and T3 as shown in Figure 4 is thus expected. By extension, we expect that specific markers elevated/decreased due to AMI onset or progression could show observable trajectories as well. Thus, we determined that Cu and Se could be more specific markers associated with the defence/response mechanism in AMI, on top of their roles in increasing general cardiovascular risk. On the other hand, significantly altered essential elements in AMI patients that did not have their levels returned closer to levels in healthy subjects after 6 months (T3) could be less specific risk factors. This applies to Mg, Na, Fe, and P. More experiments testing the levels of Cu and Se immediately after hospitalisation due to chest pain, and before/after surgical intervention may be required to understand their changes better. We discuss each significantly altered essential element in the following paragraphs in greater detail.

Firstly, for Na, the observed elevated Na levels in AMI patients (*p* < 0.05) is consistent with the literature, with the predominant view being that Na is a risk factor of CHD through increasing blood pressure and causing hypertension.^41^ Secondly, we observed lower Se levels in AMI patients (*p* < 0.01), which is also consistent with various meta-analyses on CVD and CHD.^42-44^ Available evidence indicates that the role of Se in human health and CVD is mostly attributed to its presence in antioxidant and redox-relevant selenoproteins, *e.g*., glutathione peroxidase, thioredoxin reductase.^45^ Despite that, supplementation trials did not find any significant protective effect of Se supplementation on CVD and adverse outcomes including AMI.^45^ Meanwhile, the non-significant linear association between Se and the lipid profile variables found in this work also supported such studies reporting that Se supplementation does not have effects on an aberrant lipid profile.^46^ Our work also suggests that the previous link between Se and aberrant lipid profiles could also be associated with Se’s link with Cu instead. The AMI Cu/Se levels and significant difference (*p* < 0.0001) observed in this work is firstly in line with a previous report^15^, and secondly also showed linear associations for cholesterol and LDL-C. Additionally, with reference to Table 4, we observed a significant increase in Se levels from a median of 90.31 to 94.11 ng/mL in AMI patients at T2, which remained consistent at T3. However, Se concentration at T3 (median = 94.50 ng/mL) is still significantly lower than in healthy patients (median 99.98 ng/mL; *p <* 0.05). Thus, we postulate that the lower Se levels at T1 could be related to the cardiac surgery (PCI), which may have served as an ischemic stimulus that activated inflammatory responses, thereby aggravating the decrease of circulating Se intra/-post-operation.^47, 48^

Besides Se and Na, the other significantly altered essential elements (Cu, Fe, and P) seemed to contradict the predominant views based on earlier research in cardiovascular health. In contrast, our results augment the standpoint that essential elements should have ambivalent associations with disease risk via a U- or J-shaped trend. This U/J-shaped trend is commonly associated with the dose-response relationship concept in toxicology, but it also applies to essential substances such as vitamins as well as essential trace elements.^49^ While overload/overexposure of these elements may be toxic and incur health issues, the deficiency of these essential elements may also cause adverse effects.^49^ Our results may suggest a need to redefine the normal range for these essential elements, and that the predictive value of these circulating element levels for AMI may be improved by accounting for the U/J-shaped trend.

Firstly, for Cu, Cu depletion/deficiency was more commonly reported as a risk factor of the CVDs and even a leading cause of CHD particularly.^50-52^ Cu plays essential roles in many biological processes (*e.g*., co-factor for many antioxidant enzymes, involvement in glycation, possible involvement in cytochrome and mitochondria activity, regulation of blood vessels’ response to inflammatory stimuli), and its deficiency could dysregulate these processes, leading to the promotion of disease.^50, 51^ However, there has also been contradicting reports on elevated circulating Cu found in CHD patients in large-scale studies from the USA and Finland, as well as in a smaller study in Iran.^53-56^ The results from our study agree with these reports (Table 3). Interestingly, when comparing between these studies as well as present work, it seemed that the elevated Cu was consistently seen in both prospective and cross-sectional studies.^53-56^ This indicates that Cu concentrations were elevated in CHD patients when measured both before and after atherosclerotic events. This allows us to conclude that elevated circulating Cu is likely a causal/risk factor. Nevertheless, we cannot exclude the possibility that elevated Cu is also a consequence of atherosclerotic events, as a previous study monitoring serum Cu concentrations of AMI patients over 14 days after infarct reported that serum Cu levels gradually attained peak value on the 7^th^ day after an initial rise on day 1 without returning to normal levels after 2 weeks.^57^ With reference to Table 4 and Figure 2, our study also showed that Cu levels remained elevated in AMI patients at T1 (24-48 hours after PCI) and T2 (1-month after T1), but significantly falls during the 6-month follow-up (*p* < 0.0001). While the effect size was small (W = 0.10), the result still demonstrates that the observed rise in Cu could be in part a specific defence mechanism in response to myocardial damage.^57^ Thus, we conclude that Cu could be associated with AMI as both a cause and a consequence.

Secondly, for Fe, it was reported that the conventional iron hypothesis supported by various studies was first proposed by Sullivan, that iron depletion protects against CHD.^58^ Though there were also reports with contradicting conclusions. However, it was more recently recognised that both Fe overload and deficiency are associated with increased cardiovascular morbidity and mortality.^59^ In the Ludwigshafen Risk and Cardiovascular Healthy (LURIC) study, a J-shaped association of Fe load was observed and proposed.^60^ Our work, showing lower Fe levels in AMI (*p* < 0.01), adds to the overall discussion regarding this ambivalent relationship between Fe and disease. Additionally, despite both Cu and Fe possibly having ambivalent relationships with AMI, such ambivalence does not necessarily mask the association of Fe/Cu ratio with AMI. In contrast, our results showed that Fe/Cu may be a more sensitive risk marker than either Cu or Fe alone (p < 0.0001; Table 3). However, while numerous Fe-Cu interactions were uncovered over recent years, it is unclear how the CVDs and AMI are affected. Nevertheless, the potential of Fe/Cu ratio as a sensitive marker is further revealed via multivariate analysis and classification modelling in *section 3.4* below.

Finally, for P, high phosphate levels (*i.e*., hyperphosphatemia) has been consistently linked with cardiac calcification, left ventricular hypertrophy, and cardiovascular events and death, regardless of comorbidity with chronic kidney disease (CKD).^61-63^ Concerning AMI in particular, the pro-atherogenic mechanism and formation of atherosclerotic plaques caused by phosphate excess is still unclarified and controversial.^63^ However, instead of being cardio-protective, our work shows that P levels are significantly lower in AMI patients as compared to the healthy controls (*p <* 0.05; Table 3). Our observations validate the findings by Hayward et al., who reportedly was the first to identify that low P level is also a risk factor for heart disease in adults in a large-scale UK-wide retrospective cohort study.^64^ Next, an approach using Mg supplementation had been introduced to target hyperphosphatemia-associated cardiovascular risk in CKD patients by Sakaguchi and coworkers.^21, 65^ As such, to indirectly investigate the possible balance between Mg and P with AMI and whether increased Mg levels is linked with lowered hyperphosphatemia-linked AMI, we compared the P/Mg ratios. However, our study showed that AMI patients have a significantly lower P/Mg ratio than the healthy (*p* < 0.001; Table 3). This contradicts previous findings by Sakaguchi and coworkers, suggesting that the high P and low Mg correlation could be specific to CKD patients. We thus suggest that further large-scale studies perform subgroup analysis of AMI patients with and without CKD when assessing the P and Mg relationship. A more nuanced approach is also needed when considering Mg supplementation for reducing cardiovascular mortality in CKD patients, or vice versa. Finally, besides Mg, Ca has also been linked with P, and a deranged Ca-P metabolism was identified as a major non-traditional cardiovascular risk factor for both CKD patients and even among individuals with intact renal function.^66, 67^ The link between Ca and P is multi-pronged, including Ca’s involvement in the metabolic regulation of phosphate, high phosphate contribution to vascular calcification, and valve calcification via pathological and concurrent depositions of Ca-P salts on cardiac valves.^63, 68^ However, in the current study, there is no significant association of circulating Ca levels with AMI cases (*p* > 0.05; Table 3), and we saw no significant difference in Ca×P products between groups after adjustments for covariates. This is contrary to previous results regarding coronary artery calcification and coronary heart disease risk.^13, 14^ Thus, while our study shows that Ca and P levels are indeed correlated (Figure 1), we can only conclude Ca’s involvement in phosphate metabolism (or vice versa). On the other hand, the utility of the Ca×P product as a CVD or AMI biomarker is limited and requires re-assessment.

#### 3.3.2. Non-essential or toxic elements, except As, are not specifically associated with AMI

Through the Mann Whitney test, Al, Ba, Rb, and Sr were found to be significantly elevated in AMI patients, while As levels were lowered. However, for most of them (Al, Rb, and St), their *p*-values were attenuated to non-significance upon ANCOVA. This was expected, as smoking status is a confounder for the various elements (discussed in more detail in *section 3.2*). Based on our findings, we suggest future studies definitively include smoking as a covariate for these elements to avoid inflating their roles in diseases.

Next, as for Ba, its levels were significantly higher (adjusted *p* < 0.05) in AMI patients. Ba has been seldom associated with AMI, and the few epidemiological studies assessing its link to the CVDs (especially from drinking water) did not reveal Ba as a risk factor of clinical significance.^69-71^ The reported cardiovascular effects by Ba include adverse effects on heart rhythm and hypotension, which were suggested to be caused by Ba-induced hypokalemia.^72^ However, correlation analysis did not reveal significant linear associations between Ba and K in our study populations (Figure 1). Moreover, the *p*-value from the univariate comparison between AMI and healthy was attenuated close to borderline significance after adjustments for race and white blood cell levels. Thus, since our study lack information regarding the level of exposure to Ba through any possible routes (*e.g*., drinking water or clinical sources like Ba contrast solution used in radiologic examinations), we could not make further assessments regarding the specific role of Ba in AMI. Regardless, Ba remains to be of limited significance as a risk factor for AMI.

For As, unexpectedly, we found lower As levels in AMI patients than healthy, which *p*-value remained significant after ANCOVA (*p* < 0.001). There is ample evidence that CHD risk is positively associated with inorganic As exposure (via drinking water, rice, etc).^73-76^ Other than the use of As trioxide as a chemotherapeutic or antineoplastic drug, As does not have any known benefits to human health.^77^ Since history of malignancy within the last 12-months was an exclusion criterion for the study’s sampling design, we may exclude As bioaccumulation from such exposures as a possible explanation. Alternatively, the lowered As could be an indirect response to effects induced by the PCI. This is supported by our observation that As levels at T2 and T3 were significantly increased from T1 based on the repeated measures ANOVA and post-hoc pairwise comparisons (*p* < 0.0001), and that levels at T3 were not significantly different from healthy based on a Mann Whitney test (*p* > 0.05). Further studies on As metabolism in AMI onset/intervention strategies are needed to evaluate this possibility. Regardless, the non-significant difference between As levels in AMI patients and T3 versus in healthy subjects could be attributed to the biospecimen being studied. While chronic low As exposure may result in As accumulation in blood, there are notable differences between the profiles of As species in the plasma and the urine, and these different species could be indicators of sources of As (organic forms from dietary sources, or inorganic forms from environmental exposure).^78^ Additionally, methylated forms of As have been suggested as indicators of effective biotransformation/detoxification of the inorganic As species. Other organic species such as As-betaine, which are principally introduced from seafood and agricultural products, also has low toxicity despite being found in human serum.^79^ Finally, previous work also reported different As speciation profiles in different cardiovascular tissues of CHD patients.^80^ Thus, we recommend future work to perform As speciation analysis instead of mere total As measurements in circulation, to more accurately define the contribution of As in AMI.

### 3.4. Classification modelling and performance of metallomics as AMI biomarker

Notwithstanding the possible diverse roles of the various elements as either non-specific and non-independent risk factors or as more specific markers related to cardiac injury and surgery, we performed classification modelling using all metallomic features to assess the performance of metallomics as a biomarker. Age and the traditional cardiovascular risk factors measured in routine tests (DBP, SBP, triglyceride, cholesterol, HDL-C, LDL-C, WBC, platelet, creatinine) were also included in the modelling for comparison purposes. A total of 40 variables (30 metallomic features + 10 clinical variables) was explored. The ROC curves based on models formed with varying numbers of features were generated, and predictive accuracies were also calculated as an average based on 30 iterations (Figure 3).

**Figure 3.**
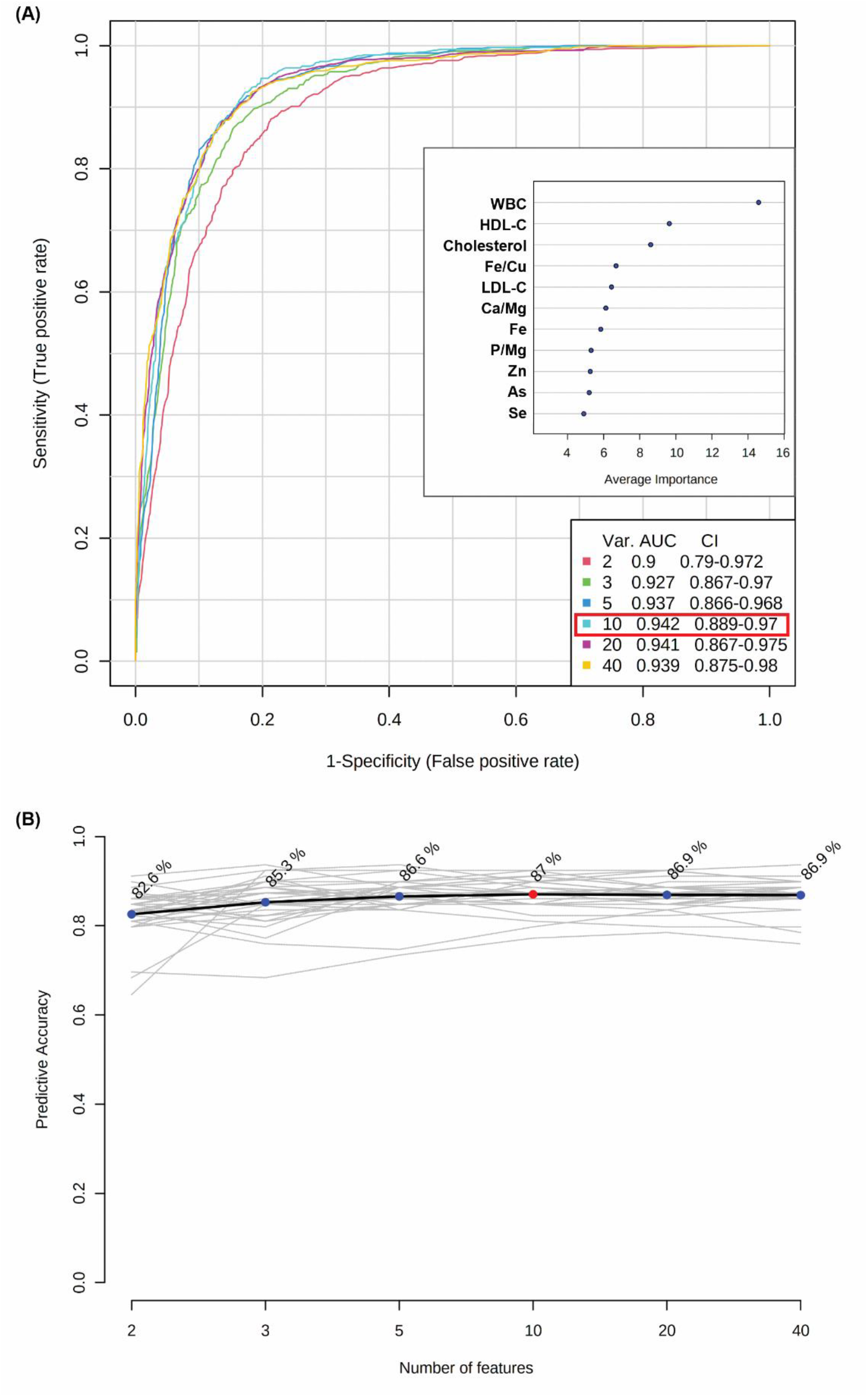
Classification performance of random forest models built with metallomic features and traditional cardiovascular risk factors as shown by (A) ROC curves and most important variables identified in the model, and (B) predictive accuracies.

As reflected in Figures 3A and 3B, the best performing model with an area under curve (AUC) of 0.942 (95% CI 0.889 - 0.970) and predictive accuracy of 87% was made up of a combination of 10 variables. The top 10 important features selected for the random forest classifier are thus also shown in Figure 3A. While the top 3 variables are clinical variables (WBC, HDL-C, and cholesterol), the following variables are all metallomic features (Fe/Cu, Ca/Mg, Fe, P/Mg, Zn, As, Se). This suggests the potential of metallomic features for the replacement or concurrent usage with current risk factors in predicting AMI.

Next, the identification of Fe/Cu ratio as an important variable augmented the univariate results in suggesting it as a potentially more sensitive biomarker for AMI than the 2 metals individually. Also, the inclusion of Cu, Se, and As as important features may also confer specificity of the model for AMI prediction or prognosis, based on postulations regarding their changes at T2 and T3 as discussed earlier. However, out of the important variables, Ca/Mg and Zn were not found to be significantly altered in AMI based on univariate comparisons. In line with random forest classifiers’ non-linear nature, our results suggest that Ca/Mg and Zn’s relationships with AMI, at least in the populations studied, could be non-linear.

## 4. Conclusions

In summary, a total of 30 metallomic features (12 essential elements, 10 non-essential or toxic elements, as well as 8 element-pair product/ratios) were profiled via a validated and efficient ICP-MS/MS-based workflow. Greater insights into the mechanisms and ambivalent relationships of the essential elements (*e.g*., Cu, Fe, and P) with health and disease were revealed based on our results and comparisons with literature. Additionally, changes in the AMI metallomic profile across 3 time-points revealed possible roles of Fe, Na, and P as stable but non-specific risk factors of AMI, and Cu and Se as more specific biomarkers. Based on the analysis of product/ratios calculated from related essential element pairs, more sensitive variables are identified (*e.g*.,Cu/Se, Fe/Cu). Classification modelling also revealed several metallomic features as potential biomarkers that may be used in replacement of or alongside traditional cardiovascular risk factors for AMI prediction.

Finally, our study has some limitations. Firstly, the sample size was relatively small, and the proposed U/J-shaped trend for the essential elements’ ambivalent associations could not be tangibly visualised. Secondly, while the longitudinal analysis across 3 time-points provided intriguing hypotheses on element concentrations’ causal/consequential links to AMI, further studies over longer periods and also specifically on closer time-points before and after intervention are required to confirm the hypotheses. Speciation analysis, especially for elements such as Se and As, are also recommended to better understand their modes of action in AMI for more specific therapeutic strategies and biomarker development.

## Data Availability

All data produced in the present study are available upon reasonable request to the authors.

## Supplementary Materials

Supplementary Information (.docx)

Table S1: Instrument settings for the Agilent 8900 ICP-QQQ in different CRC modes. Table S2: Mass shifts, ISTD, and CRC modes used for the metallomics analysis.

## Acknowledgments

The authors would like to thank the engineers and product specialists from Agilent Technologies Singapore Pte Ltd for the technical support and advice.

## References

1. Douglas P. Zipes, P. L., Robert O. Bonow, Douglas L. Mann, Gordon F. Tomaselli, Braunwald’s Heart Disease: A Textbook of Cardiovascular Medicine. 11 ed.; Elsevier: 2018.

2. Global, regional, and national age-sex-specific mortality for 282 causes of death in 195 countries and territories, 1980–2017: a systematic analysis for the Global Burden of Disease Study 2017. Lancet 2018; 392: 1736–88 2018, 392 (10159), 1736–88.

3. Chowdhury, R.; Ramond, A.; O’Keeffe, L. M.; Shahzad, S.; Kunutsor, S. K.; Muka, T.; Gregson, J.; Willeit, P.; Warnakula, S.; Khan, H.; Chowdhury, S.; Gobin, R.; Franco, O. H.; Di Angelantonio, E., Environmental toxic metal contaminants and risk of cardiovascular disease: systematic review and meta-analysis. BMJ 2018, k3310.

4. Edet Ekpenyong, C., Essential Trace Element and Mineral Deficiencies and Cardiovascular Diseases: Facts and Controversies. International Journal of Nutrition and Food Sciences 2017, 6 (2), 53.

5. Lim, S. Y.; Selvaraji, S.; Lau, H.; Li, S. F. Y., Application of omics beyond the central dogma in coronary heart disease research: A bibliometric study and literature review. Computers in Biology and Medicine 2022, 140, 105069.

6. Maret, W., An Appraisal of the Field of Metallomics and the Roles of Metal Ions in Biochemistry and Cell Signaling. Applied Sciences 2021, 11 (22).

7. Chen, B.; Hu, L.; He, B.; Luan, T.; Jiang, G., Environmetallomics: Systematically investigating metals in environmentally relevant media. TrAC Trends in Analytical Chemistry 2020, 126.

8. Walkner, C.; Gratzer, R.; Meisel, T.; Bokhari, S. N. H., Multi-element analysis of crude oils using ICP-QQQ-MS. Organic Geochemistry 2017, 103, 22–30.

9. Paseka, R. E.; Bratt, A. R.; MacNeill, K. L.; Burian, A.; See, C. R., Elemental Ratios Link Environmental Change and Human Health. Frontiers in Ecology and Evolution 2019, 7.

10. Dai, Q.; Shu, X.-O.; Deng, X.; Xiang, Y.-B.; Li, H.; Yang, G.; Shrubsole, M. J.; Ji, B.; Cai, H.; Chow, W.-H.; Gao, Y.-T.; Zheng, W., Modifying effect of calcium/magnesium intake ratio and mortality: a population-based cohort study. BMJ Open 2013, 3 (2), e002111.

11. Hibler, E. A.; Zhu, X.; Shrubsole, M. J.; Hou, L.; Dai, Q., Physical activity, dietary calcium to magnesium intake and mortality in the National Health and Examination Survey 1999–2006 cohort. International Journal of Cancer 2020, 146 (11), 2979–2986.

12. Pereira, D. D. C.; Lima, R. P. A.; De Lima, R. T.; Gonçalves, M. D. C. R.; De Morais, L. C. S. L.; Franceschini, S. D. C. C.; Filizola, R. G.; De Moraes, R. M.; Asciutti, L. S. R.; Costa, M. J. D. C., Association between obesity and calcium:phosphorus ratio in the habitual diets of adults in a city of Northeastern Brazil: an epidemiological study. Nutrition Journal 2013, 12 (1), 90.

13. Kwak, S. M.; Kim, J. S.; Choi, Y.; Chang, Y.; Kwon, M. J.; Jung, J. G.; Jeong, C.; Ahn, J.; Kim, H. S.; Shin, H.; Ryu, S., Dietary intake of calcium and phosphorus and serum concentration in relation to the risk of coronary artery calcification in asymptomatic adults. Arterioscler Thromb Vasc Biol 2014, 34 (8), 1763–9.

14. Foley, R. N.; Collins, A. J.; Ishani, A.; Kalra, P. A., Calcium-phosphate levels and cardiovascular disease in community-dwelling adults: the Atherosclerosis Risk in Communities (ARIC) Study. Am Heart J 2008, 156 (3), 556–63.

15. Mironczuk, A.; Kapica-Topczewska, K.; Socha, K.; Soroczynska, J.; Jamiolkowski, J.; Kulakowska, A.; Kochanowicz, J., Selenium, Copper, Zinc Concentrations and Cu/Zn, Cu/Se Molar Ratios in the Serum of Patients with Acute Ischemic Stroke in Northeastern Poland-A New Insight into Stroke Pathophysiology. Nutrients 2021, 13 (7).

16. Malavolta, M.; Giacconi, R.; Piacenza, F.; Santarelli, L.; Cipriano, C.; Costarelli, L.; Tesei, S.; Pierpaoli, S.; Basso, A.; Galeazzi, R.; Lattanzio, F.; Mocchegiani, E., Plasma copper/zinc ratio: an inflammatory/nutritional biomarker as predictor of all-cause mortality in elderly population. Biogerontology 2010, 11 (3), 309–319.

17. Malavolta, M.; Piacenza, F.; Basso, A.; Giacconi, R.; Costarelli, L.; Mocchegiani, E., Serum copper to zinc ratio: Relationship with aging and health status. Mech Ageing Dev 2015, 151, 93–100.

18. Collins, J. F.; Prohaska, J. R.; Knutson, M. D., Metabolic crossroads of iron and copper. Nutrition Reviews 2010, 68 (3), 133–147.

19. Cook, N. R., Joint Effects of Sodium and Potassium Intake on Subsequent Cardiovascular Disease. Archives of Internal Medicine 2009, 169 (1), 32.

20. Baer, D. J.; Althouse, A.; Hermann, M.; Johnson, J.; Maki, K. C.; Marklund, M.; Vogt, L.; Wesson, D.; Stallings, V. A., Targeting the Dietary Na:K Ratio—Considerations for Design of an Intervention Study to Impact Blood Pressure. Advances in Nutrition 2022, 13 (1), 225–233.

21. Sakaguchi, Y.; Fujii, N.; Shoji, T.; Hayashi, T.; Rakugi, H.; Iseki, K.; Tsubakihara, Y.; Isaka, Y., Magnesium Modifies the Cardiovascular Mortality Risk Associated with Hyperphosphatemia in Patients Undergoing Hemodialysis: A Cohort Study. PLoS ONE 2014, 9 (12), e116273.

22. Tanvir, E. M.; Komarova, T.; Comino, E.; Sumner, R.; Whitfield, K. M.; Shaw, P. N., Effects of storage conditions on the stability and distribution of clinical trace elements in whole blood and plasma: Application of ICP-MS. J Trace Elem Med Biol 2021, 68, 126804.

23. Robin, X.; Turck, N.; Hainard, A.; Tiberti, N.; Lisacek, F.; Sanchez, J.-C.; Müller, M., pROC: an open-source package for R and S+ to analyze and compare ROC curves. BMC Bioinformatics 2011, 12 (1), 77.

24. Konz, T.; Migliavacca, E.; Dayon, L.; Bowman, G.; Oikonomidi, A.; Popp, J.; Rezzi, S., ICP-MS/MS-Based Ionomics: A Validated Methodology to Investigate the Biological Variability of the Human Ionome. Journal of Proteome Research 2017, 16 (5), 2080–2090.

25. Meyer, S.; Markova, M.; Pohl, G.; Marschall, T. A.; Pivovarova, O.; Pfeiffer, A. F. H.; Schwerdtle, T., Development, validation and application of an ICP-MS/MS method to quantify minerals and (ultra-)trace elements in human serum. J Trace Elem Med Biol 2018, 49, 157–163.

26. McShane, W. J.; Pappas, R. S.; Wilson-McElprang, V.; Paschal, D., A rugged and transferable method for determining blood cadmium, mercury, and lead with inductively coupled plasma-mass spectrometry. Spectrochimica Acta Part B: Atomic Spectroscopy 2008, 63 (6), 638–644.

27. Flores, E. M. M.; Mello, P. A.; Krzyzaniak, S. R.; Cauduro, V. H.; Picoloto, R. S., Challenges and trends for halogen determination by inductively coupled plasma mass spectrometry: A review. Rapid Commun Mass Spectrom 2020, 34 Suppl 3, e8727.

28. Bernhard, D.; Rossmann, A.; Wick, G., Metals in cigarette smoke. IUBMB Life (International Union of Biochemistry and Molecular Biology: Life) 2005, 57 (12), 805–809.

29. Bernhard, D.; Rossmann, A.; Henderson, B.; Kind, M.; Seubert, A.; Wick, G., Increased Serum Cadmium and Strontium Levels in Young Smokers. Arteriosclerosis, Thrombosis, and Vascular Biology 2006, 26 (4), 833–838.

30. Nechifor, M., Magnesium and Zinc Involvement in Tobacco Addiction. Journal of Addiction Research & Therapy 2012, 01 (S2).

31. Richter, P.; Faroon, O.; Pappas, R. S., Cadmium and Cadmium/Zinc Ratios and Tobacco-Related Morbidities. International Journal of Environmental Research and Public Health 2017, 14 (10), 1154.

32. Suarez-Varela, M. M.; Llopis-González, A.; González Albert, V.; López-Izquierdo, R.; González-Manzano, I.; Cháves, J.; Biosca, V. H.; Martin-Escudero, J. C., Zinc and smoking habits in the setting of hypertension in a Spanish populations. Hypertension Research 2015, 38 (2), 149–154.

33. L, J.; Lutsenko, S., The Role of Copper as a Modifier of Lipid Metabolism. InTech: 2013.

34. Wagner, P.; Heinecke, J. W., Copper Ions Promote Peroxidation of Low Density Lipoprotein Lipid by Binding to Histidine Residues of Apolipoprotein B100, But They Are Reduced at Other Sites on LDL. Arteriosclerosis, Thrombosis, and Vascular Biology 1997, 17 (11), 3338–3346.

35. Holvoet, P.; Mertens, A.; Verhamme, P.; Bogaerts, K.; Beyens, G.; Verhaeghe, R.; Collen, D. S.; Muls, E.; Van De Werf, F., Circulating Oxidized LDL Is a Useful Marker for Identifying Patients With Coronary Artery Disease. Arteriosclerosis, Thrombosis, and Vascular Biology 2001, 21 (5), 844–848.

36. Islam, L.; Nabi, A.; Rahman, M.; Khan, M.; Kazi, A., Association of Clinical Complications with Nutritional Status and the Prevalence of Leukopenia among Arsenic Patients in Bangladesh. International Journal of Environmental Research and Public Health 2004, 1 (2), 74–82.

37. Zarei, M. H.; Pourahmad, J.; Nassireslami, E., Toxicity of arsenic on isolated human lymphocytes: The key role of cytokines and intracellular calcium enhancement in arsenic-induced cell death. Main Group Metal Chemistry 2019, 42 (1), 125–134.

38. Lee, C. D.; Folsom, A. R.; Nieto, F. J.; Chambless, L. E.; Shahar, E.; Wolfe, D. A., White Blood Cell Count and Incidence of Coronary Heart Disease and Ischemic Stroke and Mortality from Cardiovascular Disease in African-American and White Men and Women: Atherosclerosis Risk in Communities Study. American Journal of Epidemiology 2001, 154 (8), 758–764.

39. Batista, M. N.; Cuppari, L.; De Fátima Campos Pedrosa, L.; Almeida, M. D. G.; De Almeida, J. B.; De Medeiros, A. C. Q.; Canziani, M. E. F., Effect of End-Stage Renal Disease and Diabetes on Zinc and Copper Status. Biological Trace Element Research 2006, 112 (1), 1–12.

40. Jeremias, A., The utility of troponin measurement to detect myocardial infarction: review of the current findings. Vascular Health and Risk Management 2010, 691.

41. O’Donnell, M.; Mente, A.; Yusuf, S., Sodium Intake and Cardiovascular Health. Circulation Research 2015, 116 (6), 1046–1057.

42. Zhang, X.; Liu, C.; Guo, J.; Song, Y., Selenium status and cardiovascular diseases: meta-analysis of prospective observational studies and randomized controlled trials. European Journal of Clinical Nutrition 2016, 70 (2), 162–169.

43. Flores-Mateo, G.; Navas-Acien, A.; Pastor-Barriuso, R.; Guallar, E., Selenium and coronary heart disease: a meta-analysis. The American Journal of Clinical Nutrition 2006, 84 (4), 762–773.

44. Kuria, A.; Tian, H.; Li, M.; Wang, Y.; Aaseth, J. O.; Zang, J.; Cao, Y., Selenium status in the body and cardiovascular disease: a systematic review and meta-analysis. Critical Reviews in Food Science and Nutrition 2021, 61 (21), 3616–3625.

45. Benstoem, C.; Goetzenich, A.; Kraemer, S.; Borosch, S.; Manzanares, W.; Hardy, G.; Stoppe, C., Selenium and Its Supplementation in Cardiovascular Disease—What do We Know? Nutrients 2015, 7 (5), 3094–3118.

46. Ju, W.; Li, X.; Li, Z.; Wu, G. R.; Fu, X. F.; Yang, X. M.; Zhang, X. Q.; Gao, X. B., The effect of selenium supplementation on coronary heart disease: A systematic review and meta-analysis of randomized controlled trials. Journal of Trace Elements in Medicine and Biology 2017, 44, 8–16.

47. Koszta, G.; Kacska, Z.; Szatmári, K.; Szerafin, T.; Fülesdi, B., Lower whole blood selenium level is associated with higher operative risk and mortality following cardiac surgery. Journal of Anesthesia 2012, 26 (6), 812–821.

48. Stoppe, C.; Schälte, G.; Rossaint, R.; Coburn, M.; Graf, B.; Spillner, J.; Marx, G.; Rex, S., The intraoperative decrease of selenium is associated with the postoperative development of multiorgan dysfunction in cardiac surgical patients*. Critical Care Medicine 2011, 39 (8).

49. Moffett, D. B.; Mumtaz, M. M.; Sullivan, D. W.; Fowler, B. A., Chapter 10 - General Considerations of Dose-Effect and Dose-Response Relationships∗. In Handbook on the Toxicology of Metals (Fourth Edition), Nordberg, G. F.; Fowler, B. A.; Nordberg, M., Eds. Academic Press: San Diego, 2015; pp 197–212.

50. Dinicolantonio, J. J.; Mangan, D.; O’Keefe, J. H., Copper deficiency may be a leading cause of ischaemic heart disease. Open Heart 2018, 5 (2), e000784.

51. Klevay, L. M., Cardiovascular Disease from Copper Deficiency—A History. The Journal of Nutrition 2000, 130 (2), 489S–492S.

52. Liu, Y.; Miao, J., An Emerging Role of Defective Copper Metabolism in Heart Disease. Nutrients 2022, 14 (3).

53. Bagheri, B.; Akbari, N.; Tabiban, S.; Habibi, V.; Mokhberi, V., Serum level of copper in patients with coronary artery disease. Niger Med J 2015, 56 (1), 39–42.

54. Ford, E. S., Serum Copper Concentration and Coronary Heart Disease among US Adults. American Journal of Epidemiology 2000, 151 (12), 1182–1188.

55. Kunutsor, S. K.; Dey, R. S.; Laukkanen, J. A., Circulating Serum Copper Is Associated with Atherosclerotic Cardiovascular Disease, but Not Venous Thromboembolism: A Prospective Cohort Study. Pulse 2021, 9 (3-4), 109–115.

56. Salonen, J. T.; Salonen, R.; Korpela, H.; Suntioinen, S.; Tuomilehto, J., Serum Copper and the Risk of Acute Myocardial Infarction: A Prospective Population Study in Men in Eastern Finland. American Journal of Epidemiology 1991, 134 (3), 268–276.

57. Jain, V. K.; Mohan, G., Serum zinc and copper in myocardial infarction with particular reference to prognosis. Biological Trace Element Research 1991, 31 (3), 317–322.

58. De Valk, B.; Marx, J. J. M., Iron, Atherosclerosis, and Ischemic Heart Disease. Archives of Internal Medicine 1999, 159 (14), 1542.

59. Von Eckardstein, A., Iron in Coronary Heart Disease—J-Shaped Associations and Ambivalent Relationships. Clinical Chemistry 2019, 65 (7), 821–823.

60. Grammer, T. B.; Scharnagl, H.; Dressel, A.; Kleber, M. E.; Silbernagel, G.; Pilz, S.; Tomaschitz, A.; Koenig, W.; Mueller-Myhsok, B.; März, W.; Strnad, P., Iron Metabolism, Hepcidin, and Mortality (the Ludwigshafen Risk and Cardiovascular Health Study). Clinical Chemistry 2019, 65 (7), 849–861.

61. Foley, R. N., Phosphate Levels and Cardiovascular Disease in the General Population. Clinical Journal of the American Society of Nephrology 2009, 4 (6), 1136–1139.

62. Kendrick, J.; Kestenbaum, B.; Chonchol, M., Phosphate and Cardiovascular Disease. Advances in Chronic Kidney Disease 2011, 18 (2), 113–119.

63. Zhou, C.; Shi, Z.; Ouyang, N.; Ruan, X., Hyperphosphatemia and Cardiovascular Disease. Front Cell Dev Biol 2021, 9, 644363.

64. Hayward, N.; McGovern, A.; De Lusignan, S.; Cole, N.; Hinton, W.; Jones, S., U-shaped relationship between serum phosphate and cardiovascular risk: A retrospective cohort study. PLOS ONE 2017, 12 (11), e0184774.

65. Sakaguchi, Y.; Hamano, T.; Obi, Y.; Monden, C.; Oka, T.; Yamaguchi, S.; Matsui, I.; Hashimoto, N.; Matsumoto, A.; Shimada, K.; Takabatake, Y.; Takahashi, A.; Kaimori, J.-Y.; Moriyama, T.; Yamamoto, R.; Horio, M.; Yamamoto, K.; Sugimoto, K.; Rakugi, H.; Isaka, Y., A Randomized Trial of Magnesium Oxide and Oral Carbon Adsorbent for Coronary Artery Calcification in Predialysis CKD. Journal of the American Society of Nephrology 2019, 30 (6), 1073–1085.

66. Heine, G. H.; Nangaku, M.; Fliser, D., Calcium and phosphate impact cardiovascular risk. European Heart Journal 2013, 34 (15), 1112–1121.

67. Block, G.; Port, F. K., THE CLINICAL EPIDEMIOLOGY OF CARDIOVASCULAR DISEASES IN CHRONIC KIDNEY DISEASE: Calcium Phosphate Metabolism and Cardiovascular Disease in Patients with Chronic Kidney Disease. Seminars in Dialysis 2003, 16 (2), 140–147.

68. Massera, D.; Kizer, J. R.; Dweck, M. R., Mechanisms of mitral annular calcification. Trends Cardiovasc Med 2020, 30 (5), 289–295.

69. Wones, R. G.; Stadler, B. L.; Frohman, L. A., Lack of effect of drinking water barium on cardiovascular risk factors. Environmental health perspectives 1990, 85, 355–359.

70. Kravchenko, J.; Darrah, T. H.; Miller, R. K.; Lyerly, H. K.; Vengosh, A., A review of the health impacts of barium from natural and anthropogenic exposure. Environmental Geochemistry and Health 2014, 36 (4), 797–814.

71. Nigra, A. E.; Ruiz-Hernandez, A.; Redon, J.; Navas-Acien, A.; Tellez-Plaza, M., Environmental Metals and Cardiovascular Disease in Adults: A Systematic Review Beyond Lead and Cadmium. Current Environmental Health Reports 2016, 3 (4), 416–433.

72. Choudhury, H.; Cary, R.; World Health, O.; International Programme on Chemical, S., Barium and barium compounds. World Health Organization: Geneva, 2001.

73. States, J. C.; Srivastava, S.; Chen, Y.; Barchowsky, A., Arsenic and Cardiovascular Disease. Toxicological Sciences 2009, 107 (2), 312–323.

74. Sobel, M. H.; Sanchez, T. R.; Jones, M. R.; Kaufman, J. D.; Francesconi, K. A.; Blaha, M. J.; Vaidya, D.; Shimbo, D.; Gossler, W.; Gamble, M. V.; Genkinger, J. M.; Navas-Acien, A., Rice Intake, Arsenic Exposure, and Subclinical Cardiovascular Disease Among US Adults in MESA. Journal of the American Heart Association 2020, 9 (4).

75. Moon, K. A.; Oberoi, S.; Barchowsky, A.; Chen, Y.; Guallar, E.; Nachman, K. E.; Rahman, M.; Sohel, N.; D’Ippoliti, D.; Wade, T. J.; James, K. A.; Farzan, S. F.; Karagas, M. R.; Ahsan, H.; Navas-Acien, A., A dose-response meta-analysis of chronic arsenic exposure and incident cardiovascular disease. International Journal of Epidemiology 2017, 46 (6), 1924–1939.

76. Navas-Acien, A.; Sharrett, A. R.; Silbergeld, E. K.; Schwartz, B. S.; Nachman, K. E.; Burke, T. A.; Guallar, E., Arsenic Exposure and Cardiovascular Disease: A Systematic Review of the Epidemiologic Evidence. American Journal of Epidemiology 2005, 162 (11), 1037–1049.

77. Vineetha, V. P.; Raghu, K. G., An Overview on Arsenic Trioxide-Induced Cardiotoxicity. Cardiovascular Toxicology 2019, 19 (2), 105–119.

78. Bommarito, P. A.; Beck, R.; Douillet, C.; Del Razo, L. M.; Garcia-Vargas, G.-G.; Valenzuela, O. L.; Sanchez-Peña, L. C.; Styblo, M.; Fry, R. C., Evaluation of plasma arsenicals as potential biomarkers of exposure to inorganic arsenic. Journal of Exposure Science & Environmental Epidemiology 2019, 29 (5), 718–729.

79. Popowich, A.; Zhang, Q.; Le, X. C., Arsenobetaine: the ongoing mystery. National Science Review 2016, 3 (4), 451–458.

80. Román, D.; Pizarro, I.; Rivera, L.; Cámara, C.; Palacios, M.; Gómez, M.; Solar, C., An approach to the arsenic status in cardiovascular tissues of patients with coronary heart disease. Human & Experimental Toxicology 2011, 30 (9), 1150–1164.

