## Supplementary Information for "Plasma Metallomics Reveals Potential Biomarkers and Insights into the Ambivalent Associations of Elements with Acute Myocardial Infarction"

**Supplementary Table S1.** Instrument settings for the Agilent 8900 ICP-QQQ in different CRC modes.

| Parameters | No gas mode | He mode | $O_2$ mode | $H_2$ mode |
|---|---|---|---|---|
| MS/MS mode | Single quadruple | Single quadruple | MS/MS | MS/MS |
| RF power (W) | 1550 | | | |
| Sampling depth (mm) | 9.0 | | | |
| Plasma gas flow rate (L/min) | 0.90 | | | |
| Auxiliary gas flow rate (L/min) | 0.85 | | | |
| Cell gas flow rate (mL/min) | - | 5.5 | 30% | 7.0 |

**Supplementary Table S2.** Mass shifts, ISTD, and CRC modes used for the metallomics analysis.

| **Analyte** | **Mass or mass shift** | **ISTD used** | **CRC mode** |
|---|---|---|---|
| $^{6}$Li (ISTD) | 6 | - | No gas |
| Li | 7 | $^{6}$Li | No gas |
| Be | 9 | $^{6}$Li | No gas |
| B | 11 | $^{6}$Li | No gas |
| Na | 23 | Sc (45) | He |
| Mg | 24 | Sc (45) | He |
| Al | 27 | Sc (45) | He |
| P | 31 | Sc (45 -> 61) | $O_2$ |
| S | 32 | Sc (45 -> 61) | $O_2$ |
| K | 39 | Sc (45) | He |
| Ca | 44 | Sc (45) | He |
| Sc (ISTD) | 45<br>45 -> 45<br>45 -> 61 | - | He<br>$H_2$<br>$O_2$ |
| V | 51 | Sc (45) | He |
| Cr | 52 | Sc (45) | He |
| Mn | 55 | Sc (45) | He |
| Fe | 56 | Sc (45) | He |
| Co | 59 | Sc (45) | He |
| Ni | 60 | Sc (45) | He |
| Cu | 63 | Sc (45) | He |
| Zn | 66 | Rh | He |
| Ga | 71 | Sc (45) | He |
| As | 75 | Rh | He |
| Se | 78 -> 78 | Sc (45 -> 45) | $H_2$ |
| Rb | 85 | Sc (45) | He |
| Sr | 88 | Sc (45) | He |
| Rh (ISTD) | 103 | - | He |
| Ag | 107 | Rh | He |
| Cd | 111 | Rh | He |
| Ba | 137 | Tb | He |
| Tb (ISTD) | 159 | - | He |
| U | 238 | Tb | He |
